# Lattice Radiation Therapy with Alternating Dosimetric Peaks and Valleys

**DOI:** 10.64898/2026.01.19.26344368

**Authors:** Yingjie Song, Pan Ma, Jianrong Dai

## Abstract

**Background:** Lattice radiotherapy (LRT) delivers heterogeneous dose distribution through a three-dimensional array of vertices within the tumor. It is typically applied in 1∼5 fractions for patients with large tumor volumes. However, conventional LRT generally employs only a single vertex set, which may limit the biological advantages of this technique in multi-fraction treatments.

**Purpose:** This study proposes a novel vertex arrangement strategy in LRT aimed at improving intratumoral dose homogeneity and enhancing coverage of high-dose regions through alternating irradiation of different vertex sets.

**Materials and methods:** Patients with the gross tumor volume (GTV) between 300 cm^3^ to 2000 cm^3^ who received radiotherapy treatment at our institution were considered for inclusion. An “NaCl”-type structure was employed. Two sets of vertices (“Na”-type and “Cl”-type) were distributed within the tumor volume following a face-centered cubic (FCC) close-packed pattern analogous to the NaCl crystal structure. For each of the 10 patients with large tumor volumes (range: 319.23–1649.47 cc), two plans were generated: Plan A (optimized for “Na” vertices) and Plan B (optimized for “Cl” vertices). Each plan delivered 15 Gy per fraction to the vertices. Physical doses from Plans A and B were converted to EQD_2_ (α/β = 10 for GTV, α/β = 3 for normal tissues) and summed into three composite plans: A+A, A+B, and B+B. Plan quality was assessed using generalized equivalent uniform dose (EUD), homogeneity index (HI), D_2_, D_98_, and mean normal tissue dose (D_mean_ of NT).

**Results:** The alternating composite plan (A+B) achieved significantly greater dose homogeneity compared to non-alternating plans (A+A and B+B), with a lower HI (1.23 ± 0.08 vs. 1.70 ± 0.08 and 1.70 ± 0.09, p < 0.05) and higher EUD (3.76 ± 0.38 Gy vs. 3.48 ± 0.40 Gy and 3.42 ± 0.25 Gy, p < 0.05). The low-dose metric D_98_ was also higher in A+B (4.23 ± 0.27 Gy) than in A+A (3.92 ± 0.25 Gy) and B+B (3.94 ± 0.25 Gy). No significant difference was observed in NT D_mean_ among the three composite plans.

**Conclusion:** Alternating irradiation of two geometrically complementary vertex sets significantly improves dose coverage in high-dose regions and overall dose homogeneity without increasing normal tissue toxicity and potentially enhances therapeutic efficacy in spatially fractionated radiotherapy for large tumors.

## 1 Introduction

Spatially fractionated radiation therapy (SFRT) delivers a non-uniform dose distribution characterized by alternating high- and low-dose regions within the target tumor volume^[1]∼[3]^. High-dose regions are termed “peaks,” whereas low-dose regions are referred to as “valleys”. The earliest form of SFRT, known as GRID therapy, was proposed by Kohler in 1909^[3]∼[4]^. This technique employed physical blocks to modulate high-energy X-ray fields into pencil-like beams. Compared to standard radiotherapy, this technique allows the delivery of high doses to sub-volumes of the tumor while mitigating skin toxicity associated with uniform high-dose irradiation across the entire tumor volume^[6]∼[8]^. However, although two-dimensional GRID radiotherapy successfully delivers high-dose “hot spots” to the tumor, normal tissues along the paths of these beams inevitably receive high radiation doses as well, with superficial tissues being particularly susceptible to excessive irradiation.

In 2010, Wu^[9]^ proposed extending the 2D GRID into a 3D setup. This technique uses converging photon beams or the Bragg peak of charged particles to create an array of high-dose regions (vertices) inside the tumor. These vertices are spaced apart, forming a peak-valley dose distribution. This technique is known as Lattice radiotherapy (LRT). With the advent of IMRT/VMAT and image guidance, 3D LRT enables more flexible peak–valley arrangements and better protection of skin and surrounding normal tissues. It enhances radiotherapy effectiveness by activating immune responses, bystander effects, and other anti-tumor mechanisms^[9]∼[11]^.

Although SFRT can reduce toxicity to normal tissues, some studies^[12][13]^ have indicated that its therapeutic efficacy may be inferior to conventional radiotherapy. Using mouse models, they demonstrated a positive correlation between valley dose, equivalent uniform dose (EUD), and survival. In a review of studies prior to 2022 that treated tumor-bearing animals with minibeam radiotherapy (MBRT) or microbeam radiotherapy (MRT), Fernandez-Palomo et al^[14]^. also found that valley dose was the dosimetric parameter most strongly correlated with increased life span (ILS), rather than peak dose.

It is known that cell killing in the valley regions of SFRT relies on adjacent high-dose regions, which is attributed to the unique radiobiological effects of SFRT, where the bystander effect plays a dominant role^[15-23]^. Taking bystander signaling into account, Mahmoudi et al^[24]^. proposed a model that incorporates both direct cellular damage and damage from bystander signals to determine the total cellular damage, enabling the calculation of the surviving fraction (SF) within each voxel. To further investigate the influence of bystander signals on the dose distribution in SFRT planning, their model computed a signal-adjusted dose distribution (D_sig_) for each plan and signaling range. For each voxel, the SF derived from the model was converted into an equivalent dose using the linear-quadratic (LQ) model at the same survival level, and this dose distribution was compared with the physical dose distribution. However, even after accounting for the bystander effect, the maximum effective dose in the valley regions increased by up to 150%, yet remained lower than the peak dose—in some areas only about 10% of the peak dose. Similarly, a 2025 study by Balvasi et al^[25]^., which also incorporated intercellular signaling, retrospectively selected a lung cancer patient and generated both GRID and LATTICE plans. After considering the bystander effect, the peak dose ranged between 10–14 Gy, while the valley dose remained relatively insufficient at 6–8 Gy.

Current literature on LRT shows that most LRT is combined with conventional radiotherapy, delivered in 1 to 5 fractions either before or after standard treatment. Studies by Blanco Suarez et al^[26]^., Ferini et al^[27]^., Amendola et al^[28]^., and Borzov et al^[29].^ employed 1–3 LRT sessions combined with conventional RT, with single-fraction prescription doses of 3 Gy, 15 Gy, 8 Gy, and 20 Gy per vertex, respectively. Other studies applied LRT alone: Iori et al^[30]^., Duriseti et al^[31]^., Schiff JP et al^[32]^., and Dicer et al^[32]^ . used 5-fraction LRT with vertex doses of 11 Gy, 13.34 Gy, and 10 Gy, respectively. Even in multi-fraction regimens, only a single vertex position was typically used, concentrating high doses within small volumes. Despite the indirect radiobiological benefits associated with spatially fractionated techniques, the cumulative effective dose in these low-dose regions remains relatively low, ultimately compromising overall tumor control.

Close packing is a crystallographic concept describing arrangements that maximize packing density. There are two fundamental types of close packing: face-centered cubic (FCC) and hexagonal close packing (HCP), both of which attain a maximum space utilization of 74.05%^[33]^.Previous studies^[31][34]^ have employed close-packing principles for arranging vertices in LRT.

To address the aforementioned challenges, our study aims to employ a peak-valley alternation approach by shifting the vertex positions across different fractions. The structure of sodium chloride (NaCl) crystal exhibits a unique characteristic in which both chloride and sodium ions adopt a face-centered cubic close-packed arrangement, alternating uniformly in a 1:1 ratio. We propose a novel vertex arrangement strategy inspired by the NaCl lattice. Two complementary vertex sets are distributed within the tumor according to an FCC pattern, with one set occupying sodium ion positions and the other occupying chloride ion positions, as shown in Fig. 1. Each set of vertices is designated as a peak-dose region in the treatment plan, with the assumption that only one set is targeted per fraction, alternating between the two in successive fractions. This method offers two key advantages. First, it makes full use of the biological benefits of SFRT in each fraction. Second, it improves overall tumor dose homogeneity when the plans from all fractions are combined.

**FIGURE 1.**
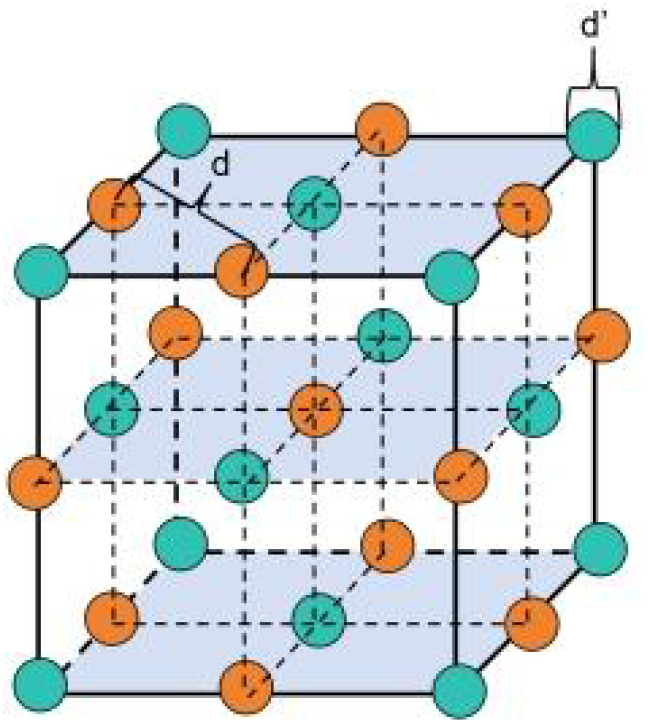
Face-centered cubic (FCC) close-packed configuration. The blue color represents the ‘Na’ ions, corresponding to the positions of the “Na”-type vertices in this study, while the orange color denotes the ‘Cl’ ions, indicating the locations of the “Cl”-type vertices. Both the size of the vertices and the center-to-center spacing are fully adjustable.

## 2 Materials and Methods

### 2.1 Patient selection

Patients who received radiotherapy treatment at our institution with the gross tumor volume (GTV) between 300 cm^3^ to 2000cm^3^ was selected, shown as Table 1. For each patient, the dataset consisted of CT images and contours of regions of interest, including target volumes and organs at risk. Ethical approval for this study was obtained from the Ethics Committee under the reference number NCC2024C-031.

**Table 1.** Patient and Tumor Characteristics (ranked from smallest to largest volume)

| Patient<br>Number | Site | Histology | GTV(cm<sup>3</sup>) |
| --- | --- | --- | --- |
| 1 | Pelvis | Malignant long bone tumor of<br>the lower extremity | 319.23 |
| 2 | Leg | Chondrosarcoma | 543.63 |
| 3 | Hip | Malignant long bone tumor of<br>the lower extremity | 658.09 |
| 4 | Leg | Soft tissue sarcoma | 789.92 |
| 5 | Abdomen | Soft tissue sarcoma | 862.05 |
| 6 | Leg | Soft tissue sarcoma | 894.79 |
| 7 | Retroperitoneum | Soft tissue sarcoma | 938.38 |
| 8 | Leg | Soft tissue sarcoma | 1407.00 |
| 9 | Abdomen | Soft tissue sarcoma | 1456.63 |
| 10 | Retroperitoneum | Peritoneal malignancy | 1649.47 |

GTV: gross tumor volume

### 2.2 Method

Vertex placement was automated using an in-house script with the following steps: 1) A lattice volume was generated by contracting the GTV inward by a defined margin. 2) Two sets of vertices were uniformly distributed based on the sodium chloride (NaCl) crystal structure, determining their central positions and spacing. 3) The size of each vertex was defined based on the percentage of the total volume of vertices relative to the volume of the GTV.

#### 2.2.1 Generation of the Lattice Volume

Because vertices deliver high doses, sufficient clearance from organs at risk (OARs) was required. By analyzing patient CT images and the contours of the target volume and organs at risk, the relative spatial relationship between normal tissues and the tumor was determined to generate the lattice volume. The lattice volume was created by contracting the GTV inward by a specific margin. The inward contraction distance (d_inward_) was derived based on fitted reference data^[2][34]^, as expressed in the following formula (Equation 1):

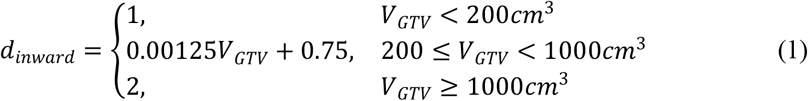

where V_GTV_ represents the volume of the patient’s GTV.

#### 2.2.2 Distribution of Two Sets of Vertex Centers Based on Sodium Chloride Crystal Structure

The crystal structure of sodium chloride exhibits a unique arrangement in which both chloride and sodium ions adopt a face-centered cubic (FCC) close-packed configuration. Following this principle, two sets of vertices—designated ‘Na-type’ and ‘Cl-type’—were distributed within the lattice volume. Both sets were identical in number, size, and overall volume.

The specific procedure is as follows:

First, we generated the first set of vertices, designated as “Na”-type vertices. The center-to-center distance d between adjacent vertices was determined based on a formula (Equation 2) derived from fitted experimental data in the literature^[2][34]^:

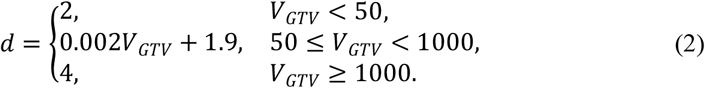

where V_GTV_ represents the volume of the patient’s GTV.

#### 2.2.3 Determination of Vertex Size

The size of the vertices is determined by the tumor volume. The diameter d’(in cm) of each vertex is calculated based on fitted data from reference literature^[2][34]^, as follows:

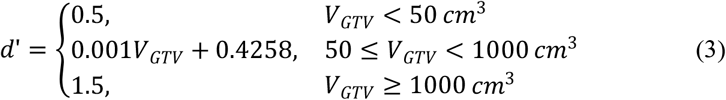

where V_GTV_ represents the volume of the patient’s GTV.

This value of d′serves as the initial diameter. Although all vertex centers were confined to the lattice volume, spherical geometry sometimes led to partial protrusion. Any volume extending beyond the lattice was removed. The total volume of each set of vertices was then calculated separately. If the total volume was less than 1% of the GTV volume, the value of d′was iteratively increased until the total volume reached or slightly exceeded 1% of the GTV volume, thereby determining the final vertex diameter.

### 2.3 Treatment Planning

Treatment planning was performed using the Pinnacle^3^ treatment planning system (version 9.16; Philips Healthcare, Andover, MA, USA). For tumors with a length less than 20 cm, plans were designed using 6 MV Flattening Filter-Free (FFF) photon beams and were intended for delivery on a Varian Edge linear accelerator (Varian Medical Systems, Palo Alto, CA, USA) equipped with a 60-pair multi-leaf collimator (MLC). The central 10 cm of the MLC had a leaf width of 2.5 mm, while the remaining leaves had a width of 5 mm. For tumors longer than 20 cm, 6 MV FFF beams were also used, with plans designed for delivery on an Elekta VersaHD linear accelerator equipped with an 80-pair MLC, all leaves of which had a width of 5 mm. The dose calculation grid resolution was set to 0.2 × 0.2 × 0.2 cm^3^. Between 3 and 5 coplanar partial arcs were used, with the exact number determined based on the geometric complexity of the tumor.

The planning objective was to deliver the prescription dose of 15 Gy to at least 95% of the volume of the vertices in a single fraction. To enforce rapid dose fall-off, three concentric ring structures were generated around each vertex: Ring 1 at a 5 mm distance from the vertex boundary, Ring 2 at 10 mm, and Ring 3 at 15 mm. While one set of vertices was designated to receive the peak dose, the dose to the other set was constrained to help reduce the dose in the valley regions. Furthermore, patient-specific optimization was performed to minimize the dose to normal tissues outside the GTV, while maintaining the maximum peak dose and minimum valley dose.

Two plans were created for each patient: Plan A was optimized with the “Na”-type vertices as the peak dose regions, and Plan B used the “Cl”-type vertices. The same dose-volume constraints were applied to all plans, and the final dose distributions were computed using adaptive convolution.

### 2.4 Treatment Plan Evaluation and Comparative Statistics

For both Plan A and Plan B, the following vertex parameters were recorded: number, diameter, center-to-center distance, and relative volume (% of GTV), as detailed in Table 2.

**Table 2.**
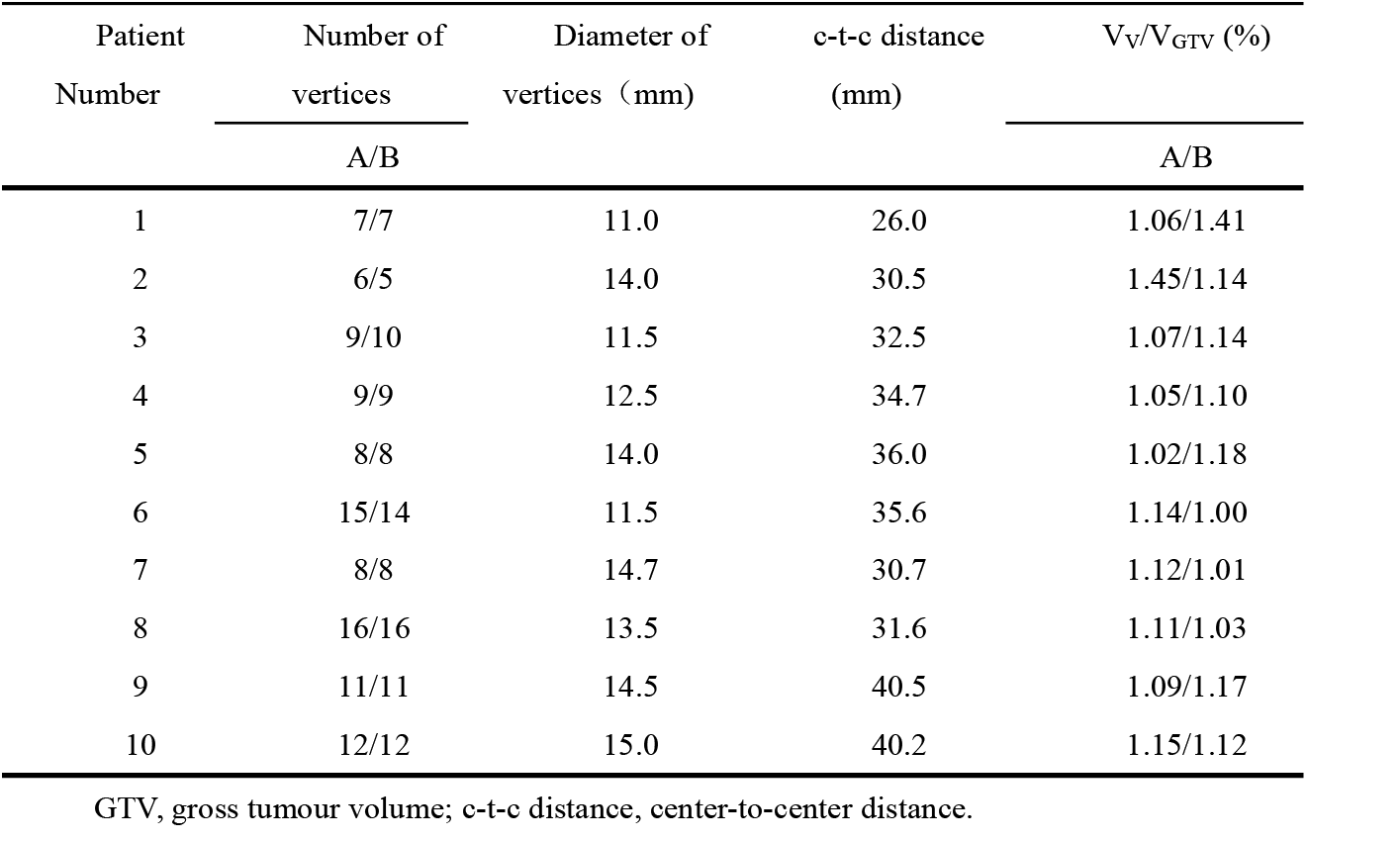
Plan A and Plan B Vertex Data Statistics.

To evaluate the proposed alternating irradiation approach, physical doses from Plan A and Plan B were converted into equivalent dose in 2-Gy fractions (EQD_^2^_) using the linear–quadratic (LQ) model:

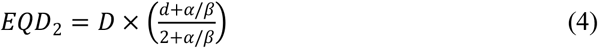

Where D is the prescription dose of 15 Gy, d is the dose of each pixel in one fraction, α/β is the tissue-specific linear-quadratic parameter. An α/β value of 10 was used for the GTV and 3 for normal tissues^[35][36]^.

Subsequently, the EQD_2_-transformed Plan A and Plan B were summed in different combinations: two fractions of Plan A were combined as Plan ‘A+A’; one fraction of Plan A and one of Plan B were summed as Plan ‘A+B’; and two fractions of Plan B formed Plan ‘B+B’.

The concept of EUD is often used to predict the effectiveness of lattice radiotherapy in terms of tumor control and normal tissue toxicity. According to Niemierko’s phenomenological model^[35][36]^, EUD is defined as:

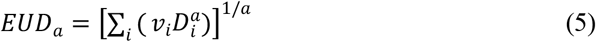

where v_i_ is the fractional subvolume receiving dose D_i_, and a is a tissue-specific scaling factor reflecting radiation sensitivity. Tumors are traditionally assigned large negative values of a; in this study, a = -10 was used^[2][37]^.

The Homogeneity Index (HI) was employed to evaluate the uniformity of the composite dose distribution. The HI formula used in this study is given by Equation (6):

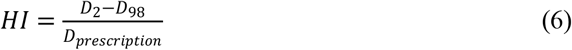

where D_2_ and D_98_ are the minimum doses delivered to 2% and 98% of the GTV volume, respectively, and D_prescription_ is the prescription dose of 15 Gy. An HI value closer to 0 indicates a more homogeneous dose distribution^[38]^.

We computed and compared the EUD and HI values for Plans ‘A+A’, ‘A+B’, and ‘B+B’. The values of D_2_ and D_98_ were used to characterize the high-dose and low-dose regions within the GTV. Normal tissue (NT) was defined on each slice by subtracting both the gross tumor volume (GTV) from the patients contour. The mean dose of NT was then used to assess toxicity in all normal tissues.

## 3 Results

### 3.1 Representative patient

Figure 2 displays the dose distribution on the same sagittal plane for a representative patient (Patient 3). The lower-left inset of each panel illustrates the corresponding vertex distribution for the different plan combinations. The patient’s GTV(red) was 629.24 cm^3^ , and the lattice volume(blue) was 221.16 cm^3^ .Cyan denotes “Na”-type vertices, and lavender represents “Cl”-type vertices. There were 10 “Na”-type vertices (Plan A) and 12 “Cl”-type vertices (Plan B), accounting for 1.07% and 1.14% of the GTV volume, with volumes of 6.73 cm^3^ and 7.20 cm^3^, respectively. The center-to-center distance of vertices was 34.7 mm, and the vertex diameter was 12.5 mm. After combination, Plan A+B resulted in a denser high-dose region within the GTV compared to Plan A+A and Plan B+B. Since vertex distribution is three-dimensional, a representative slice was selected for illustration.

**FIGURE 2.**
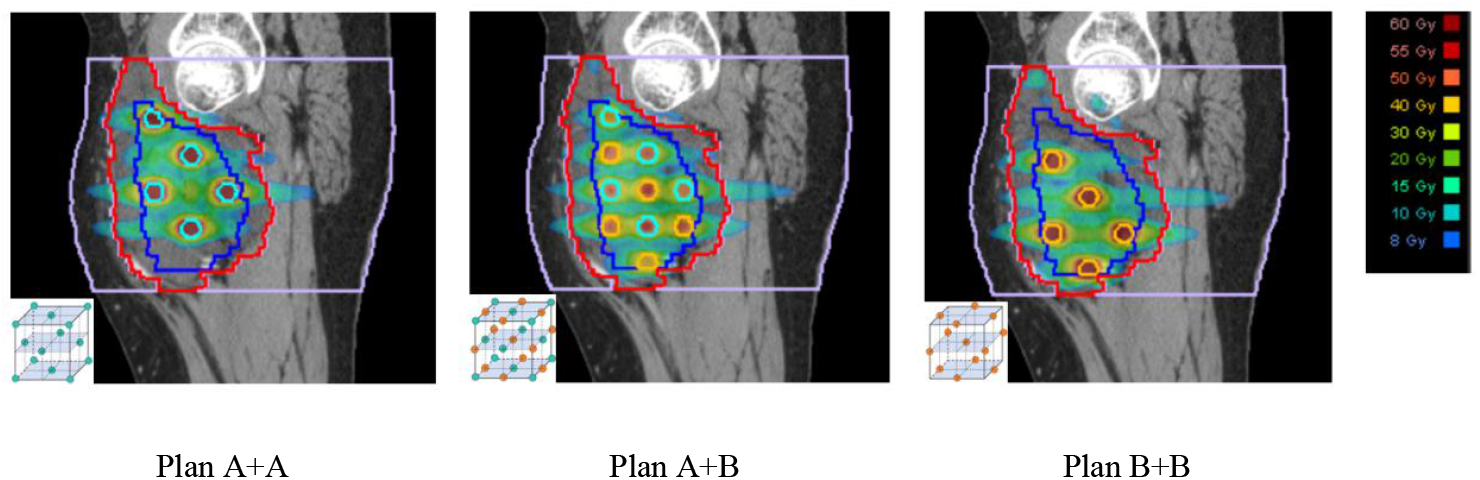
Vertex and Dose Distribution in a Representative Patient. The red area represents the gross tumor volume (GTV), while the blue region indicates the lattice volumes. Cyan denotes “Na”-type vertices, and orange represents “Cl”-type vertices. lavender corresponds to the normal tissue (NT). The lower-left inset of each panel illustrates the corresponding vertex distribution for the different plan combinations.

### 3.2 Statistics

For both Plan A and Plan B, the diameter and the center-to-center distance of vertices were consistent. The number and volume of vertices were nearly identical, with the total volume of the vertices accounting for ⩾ 1% of the GTV volume, as shown in Table 2.

After plan optimization, the volume ratio of the vertices receiving at least the prescription dose reached ⩾ 95%. After EQD_2_ conversion and dose summation of Plans A and B, the EUD, D_2_, D_98_, HI, and mean normal tissue dose (D_mean_ of NT) for the composite plans (A+A, A+B, and B+B) were calculated and are summarized.

Subsequently, appropriate statistical approaches were applied to analyze differences among the composite plans (A+A, A+B, and B+B). The specific statistical procedures were as follows: Normality of the data were first tested. If the data met normality assumptions (p > 0.05), a repeated-measures ANOVA was performed, followed by Bonferroni-adjusted pairwise comparisons. If the normality assumption was violated (p < 0.05), the non-parametric Friedman test was used, followed by Wilcoxon signed-rank tests with Bonferroni adjustment.

The results showed that for EUD, D_2_, D_98_, and HI, significant differences were observed between Plan ‘A+B’ and Plan ‘A+A’, as well as between Plan ‘A+B’ and Plan ‘B+B’, as shown in figure.3(a)∼fig.3(d). However, no significant difference was found between Plan ‘A+A’ and Plan ‘B+B’. For the mean dose to normal tissues (D_mean_), no significant differences were detected among any of the three composite plans, as show in fig.3(e).

**FIGURE 3.**
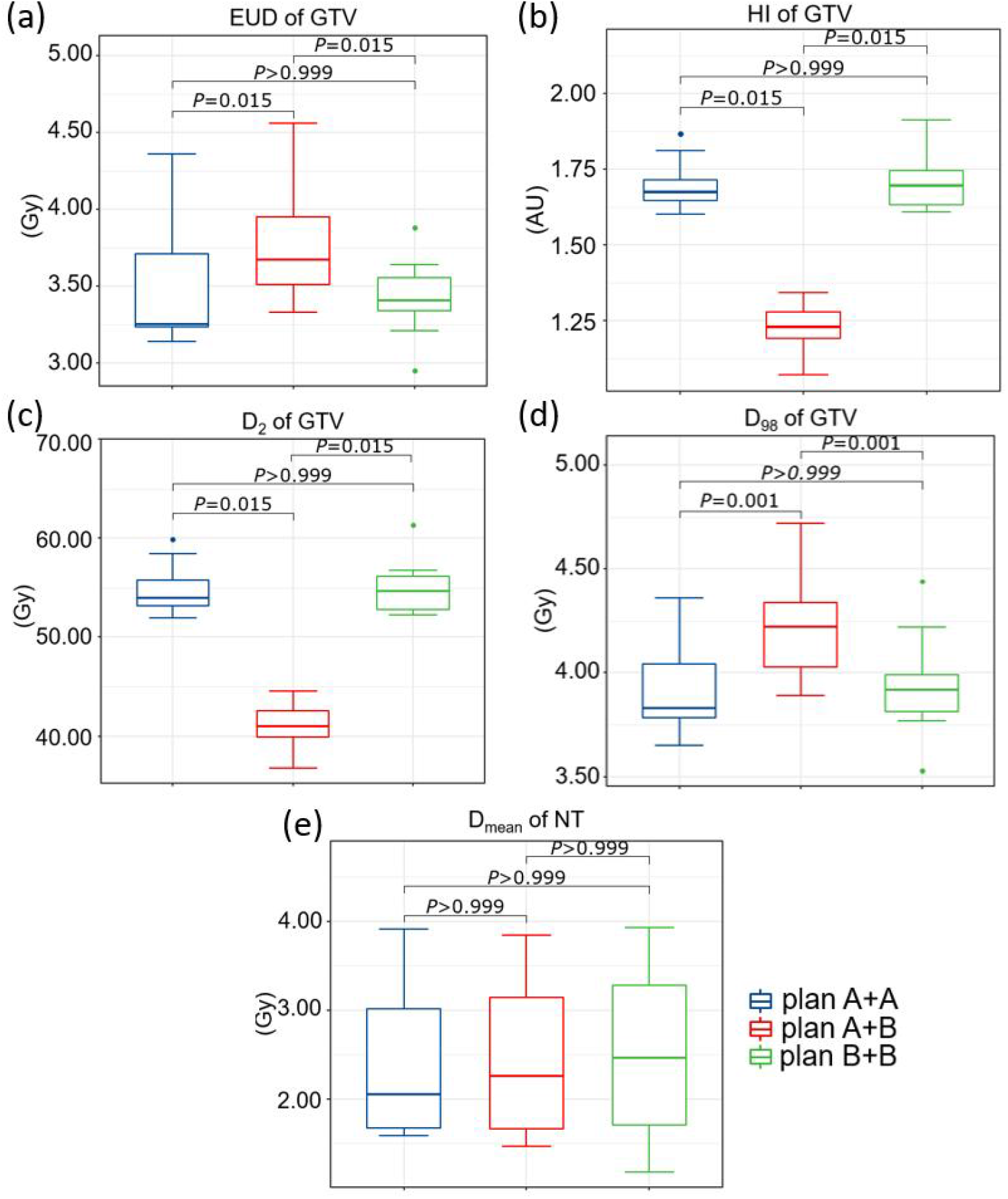
Statistical Analysis of Plan A+A Versus Plan A+B Versus Plan B+B.

Specifically, Plan ‘A+B’ achieved a higher EUD (3.76 ± 0.38 Gy) compared to Plan ‘A+A’ (3.48 ± 0.40 Gy) and Plan ‘B+B’ (3.42 ± 0.25 Gy). The low-dose metric D_98_ was also higher in Plan ‘A+B’ (4.23 ± 0.27 Gy) than in Plan ‘A+A’ (3.92 ± 0.25 Gy) and Plan ‘B+B’(3.94 ± 0.25 Gy). In contrast, the high-dose metric D_2_ was lower in Plan ‘A+B’ (41.05 ± 2.25 Gy) compared to Plan ‘A+A’ (54.90 ± 2.54 Gy) and Plan ‘B+B’ (55.03 ± 2.73 Gy). The Homogeneity Index (HI) of Plan ‘A+B’ (1.23 ± 0.08) was closer to 0 than that of Plan ‘A+A’ (1.70 ± 0.08) and Plan ‘B+B’ (1.70 ± 0.09), indicating improved dose uniformity. No significant differences were observed in the mean dose to normal tissues among Plan ‘A+A’, ‘A+B’, and ‘B+B’.

## 4 Discussion and Conclusion

For plan evaluation, we employed the EUD, widely used in radiotherapy. The parameter a plays a pivotal role in determining EUD values. While existing studies consistently use a = –10 for tumors, this may underestimate the therapeutic effect LRT, as EUD does not account for the unconventional biological mechanisms unique to SFRT^[2]^. Several potential mechanisms have been proposed, including radiation-induced bystander effects, microvascular effects, and immunomodulatory effects. These models suggest that bystander effects under non-uniform dose conditions may contribute positively to treatment efficacy, allowing a more accurate prediction of tumor control in SFRT^[15]∼[23]^. However, these models have thus far been validated only in vitro (across various cell lines) and in small-animal tumor models, and have not yet been applied in clinical practice. Furthermore, no existing model integrates all three effects simultaneously. Future work should focus on refining and validating these radiobiological models—originally developed based on preclinical data—with clinical datasets, ultimately leading to the development of a unified multi-effect model for comprehensive and accurate prediction of SFRT outcomes.

From a clinical perspective, implementation requires the creation of at least two treatment plans based on the prescribed dose and fractionation scheme for alternating delivery. Given that the biological effects of SFRT rely on heterogeneous dose distributions and require sufficient time to fully manifest, it is recommended that the interval between treatment fractions be at least 24 hours. Theoretical support for this recommendation is provided by the study of McMahon et al[39]., whose model indicates that the duration of radiation-induced bystander signal secretion t_sig_ is proportional to the absorbed dose (D), expressed as t_sig_= γ × D, where γ is a cell-type-specific parameter. For instance, in a skin model where γ is 180 min/Gy, an absorbed dose of 10 Gy would correspond to a bystander signal secretion duration of approximately 30 hours. Furthermore, in vitro experiments from the same study confirmed that as the exposure time of cells to signaling molecules in the medium increases, clonogenic survival gradually decreases and reaches saturation after about 2 hours. Therefore, to ensure that the bystander effect-mediated cell killing is fully realized—particularly as cumulative doses from multiple fractions lead to more homogeneous overall dose distribution—adequate time between SFRT fractions should be allowed. The two sets of vertices are arranged according to a sodium chloride crystal structure, classified as “Na”-type and “Cl”-type vertices. The size and spacing of the vertices should be customized according to the size and location of the patient’s tumor. Each plan uses one set of vertices (“Na” or “Cl”) as high-dose regions under identical prescription dose and optimization constraints. During treatment, the plans are delivered alternately—one set per session. If more than two plans are used, it is essential to ensure that all vertex sets are similar in size, number, and total volume, uniformly distributed, and sufficiently distant from the GTV/OAR boundaries to minimize normal tissue toxicity. All plans must also use the same prescription dose and optimization criteria^[2][4]^. While this work investigated only a two-set NaCl-inspired configuration, LRT regimens involving an odd number of fractions may necessitate extending the approach to three or more vertex sets. Future research could also focus on optimized methods for generating multi-set vertex distributions.

We use two alternating sets of targets—”Na”-type and “Cl”-type—uniformly arranged and alternately irradiated as vertices, each delivered once. Our method improves dose homogeneity within the GTV after summation compared to irradiating the same set of targets twice. This is achieved without increasing high-dose exposure or normal tissue dose. Additionally, the low-dose coverage and EUD were enhanced, all of which are beneficial for tumor control.

## Data Availability

Data are available from the corresponding author on reasonable request.

## 5 Declarations

## 5.1 Abbreviations

SFRT: spatial fractionated radiotherapy
LRT: lattice radiotherapy
MRT: microbeam radiotherapy
MBRT: minibeam radiotherapy
ILS: increased life span
SF: surviving fraction
NaCl: sodium chloride
FCC: face-centered cubic
HCP: hexagonal close packing
GTV: gross tumor volume
OAR: organ at risks
EUD: Equivalent Uniform Dose
LQ: linear-quadratic

## 5.2 Ethics approval and consent to participate

This study was conducted in accordance with the principles of the Declaration of Helsinki. The study involving human participants was reviewed and approved by the Institutional Review Board (IRB) of National Cancer Center/Cancer Hospital, Chinese Academy of Medical Sciences and Peking Union Medical College (Approval No.: NCC2024C-031). Informed consent was obtained from all individual participants. The requirement for written informed consent from participants or their legal guardians/next of kin was waived by the approving IRB, in accordance with national regulations and institutional policies. Clinical trial number: not applicable.

## 5.3 Consent for publication

Not Applicable..

## 5.4 Availability of data and materials

Data are available from the corresponding author on reasonable request.

## 5.7 Authors’ contributions

YS: Conceptualization, Investigation, Formal Analysis, Methodology, Resources, Software, Validation, Writing–original draft. PM: Data curation, Project administration, Resources, Supervision, Writing – review & editing. JD: Conceptualization, Formal Analysis, Investigation, Methodology, Project administration, Writing–review & editing. All authors have read and approved the final version of the manuscript.

## 5.8 Acknowledgements

The authors wish to thank the National Cancer Center/National Clinical Research Center for Cancer/Cancer Hospital, Chinese Academy of Medical Sciences and Peking Union Medical College for the provision of patient data and the software platform.

